# Comparing a novel neuroanimation experience to conventional therapy for high-dose, intensive upper-limb training in subacute stroke: The SMARTS2 randomized trial

**DOI:** 10.1101/2020.08.04.20152538

**Authors:** John W. Krakauer, Tomoko Kitago, Jeff Goldsmith, Omar Ahmad, Promit Roy, Joel Stein, Lauri Bishop, Kelly Casey, Belen Valladares, Michelle D. Harran, Juan Camilo Cortés, Alexander Forrence, Jing Xu, Sandra DeLuzio, Jeremia P. Held, Anne Schwarz, Levke Steiner, Mario Widmer, Kelly Jordan, Daniel Ludwig, Meghan Moore, Marlena Barbera, Isha Vora, Rachel Stockley, Pablo Celnik, Steven Zeiler, Meret Branscheidt, Gert Kwakkel, Andreas R. Luft

**Affiliations:** Dept. of Neurology, Johns Hopkins University, Baltimore, MD, USA; Dept. of Neuroscience, Johns Hopkins University, Baltimore, MD, USA; Dept. of Physical Medicine and Rehabilitation, Johns Hopkins University, Baltimore, USA; Burke Neurological Institute, White Plains, NY, USA; Dept. of Neurology, Weill Cornell Medicine, New York, NY, USA; Dept. of Neurology, Columbia University, New York, NY, USA; Dept. of Biostatistics, Columbia University Mailman School of Public Health, New York, NY, USA; Dept. of Rehabilitation and Regenerative Medicine, Columbia University Vagelos College of Physicians and Surgeons, New York, USA; cereneo Center for Neurology and Rehabilitation, Vitznau, Switzerland; School of Nursing, University of Central Lancashire, Preston, UK; Division of Vascular Neurology and Neurorehabilitation, Dept. of Neurology, University Hospital and University of Zurich, Zurich Switzerland; Amsterdam UMC, Vrije Universiteit Amsterdam, Amsterdam, Netherlands; Amsterdam Rehabilitation Research Centre, Reade, Amsterdam, Netherlands

**Keywords:** stroke, motor recovery, upper limb, neuroanimation, rehabilitation

## Abstract

**Background:** Evidence from animal studies suggests that greater reductions in post-stroke motor impairment can be attained with significantly higher doses and intensities of therapy focused on movement quality. These studies also indicate a dose-timing interaction, with more pronounced effects if high-intensity therapy is delivered in the acute/subacute, rather than chronic, post-stroke period.

**Objective:** To compare two approaches of delivering high-intensity, high-dose upper limb therapy in patients with subacute stroke: a novel exploratory neuro-animation therapy (NAT), and modified conventional occupational therapy (COT).

**Methods:** Twenty-four patients were randomized to NAT or COT and underwent 30 sessions of 60 minutes time-on-task in addition to standard care. The primary outcome was the Fugl-Meyer Upper Extremity motor score (FM-UE). Secondary outcomes included: Action Research Arm Test (ARAT), grip strength, Stroke Impact Scale (SIS) hand domain, and upper-limb kinematics. Outcomes were assessed at baseline, and days 3, 90, and 180 post-training. Both groups were compared to a matched historical cohort (HC), which received only 30 minutes of upper limb therapy per day.

**Results:** There were no significant between-group differences in FM-UE change or any of the secondary outcomes at any timepoint. Both high-dose groups showed greater recovery on the ARAT (7.3 ±2.9 pts, p=0.011), but not the FM-UE (1.4 ±2.6 pts, p =0.564) when compared to the HC.

**Conclusions:** Two forms of high-dose intensive upper limb therapy produced greater activity but not impairment improvements compared with regular care. Neuroanimation may offer a new enjoyable, efficient and scalable way to deliver increased upper limb therapy.

Clinicaltrials.gov registration NCT02292251

## INTRODUCTION

Current neurorehabilitation approaches in the subacute period after stroke have been ineffective in reducing motor impairment beyond what is expected from spontaneous biological recovery plus standard of care^1,2^. Notably, however, recent studies in patients with chronic stroke have shown improvements with promising effect sizes at both the activity and impairment levels when greatly increased intensities and doses of regular upper limb therapy are provided^3-6^.

Many studies in non-human primates, some going back over a century, have shown that hemiparesis caused by induced focal lesions in motor cortical areas and/or their descending pathways can markedly improve with high-intensity and high-dose training regimens focused on movement-quality; especially when it is initiated early (within days and weeks of the injury)^7,8^. For example, in one study, monkeys began training on day five post-infarct and were trained on 600 pellet retrievals a day for three to four weeks^9^; full recovery of hand function was seen. In another study, but this time with an emphasis on accurate reaching, a monkey began training sessions two weeks after a total arm area lesion of primary motor cortex^10^. Each session lasted 60-90 minutes, and training was given four days a week for a month. Again, the monkey attained near-normal reach performance. This amount and type of upper limb training emphasizing movement quality of the affected limb does not occur in standard rehabilitation, which instead incentivizes task accomplishment via compensation, with less emphasis on movement quality^11^. Furthermore, it has been reported that patients make a total of only about 30 upper limb task-based repetitions during a single therapy session^12^ – almost two orders of magnitude lower than in the cited monkey studies.

The goals of the SMARTS2 study were the following: 1) to test the feasibility and efficacy of upper limb therapy focused on movement quality when provided at intensities and doses comparable to those given in non-human primate studies, 2) to initiate high-intensity and high-dose therapy early, given the extensive evidence in animal models that there is increased responsiveness to training in the first weeks to a month after stroke^13^, and that most spontaneous recovery occurs in the first few months after stroke in humans^14^, 3) to devise a new immersive and enjoyable animated experience that would motivate patients to make a large number of high-quality movements in 3D, and 4) to develop outcome assessments based on movement kinematics.

SMARTS2 was a multicenter, single-blinded, parallel randomized controlled trial in subacute stroke, comparing the efficacy of intensive modified conventional occupational therapy (COT) with a new non-task-oriented approach focusing on playful exploration and quality of movement execution while controlling an immersive physics-based animated dolphin (“neuroanimation”, NAT)^15^. The hypothesis was that movement quality-focused NAT would be superior to COT at reducing impairment, as measured with the Fugl-Meyer Upper Extremity motor score (FM-UE), and at least as good at improving arm activity, measured with the Action Research Arm Test (ARAT), as we expected a large reduction in impairment to generalize to activity. The secondary hypothesis was that both forms of high-intensity and high-dose therapy would be better than standard-of-care levels of occupational therapy because there is more time to focus on movement quality.

## METHODS

### Study Design

This was a multicenter, single-blinded, parallel randomized controlled trial comparing the efficacy of a novel exploratory neuroanimation therapy (NAT) with time-matched conventional occupational therapy (COT) to enhance upper limb motor recovery after stroke. Eligible patients within six weeks post-stroke were randomized 1:1 to either NAT or COT using a central web-based database (Research Electronic Data Capture, REDCap). Randomization was blocked in groups of six and stratified according to baseline FM-UE score of 6-20 or 21-40. The randomization sequence was generated by a statistician not involved in the study and concealed from all study team members.

The target therapy schedule for both groups was two daily sessions separated by at least an hour break, five days per week for three weeks, for a total of 30 sessions. Deviations from this schedule were allowed as long as all therapy sessions could be completed within 10 weeks post-stroke. NAT and COT were matched for active therapy time of 60 minutes of time-on-task per session, which was tracked by the gaming software in the NAT group and by stopwatch in the COT group.

Outcome assessments were performed at four timepoints: baseline (pre-training), and post-training day 3 (±2 days), day 90 (±10 days), and day 180 (±10 days). All assessments were conducted by trained evaluators who were blinded to treatment allocation. Patients and caregivers were coached and given a written and verbal reminder not to speak to the evaluator regarding the therapy type at each assessment visit. The evaluators had no contact with the participants outside of the assessment sessions to minimize chances of unblinding.

The trial design initially included a third arm consisting of NAT with transcranial direct current stimulation (tDCS). Due to slow initial recruitment, the NAT + tDCS arm was stopped to allow for increased recruitment in the NAT and COT groups, which was our primary comparison of interest. The time window for enrollment post-stroke was also extended from five weeks to six weeks to increase the number of eligible participants.

### Study Participants

Patients were recruited from the acute stroke and inpatient rehabilitation units at Johns Hopkins Hospital, New York Presbyterian Hospital-Columbia, cereneo Center for Neurology and Rehabilitation, and their affiliated institutions. Patients were eligible for the study if they were 21 years old or over, had an ischemic stroke (hemispheric or brainstem) confirmed by CT or MRI within the previous six weeks with residual arm weakness (FM-UE score 6-40 pts), had no history of prior stroke with associated motor deficits, and were able to give informed consent and understand the tasks involved. Exclusion criteria included: intracranial hemorrhage, botulinum toxin injection to upper limb since stroke, physical or neurological condition that interfered with study procedures or assessment of upper limb motor function, inability to sit in a chair and exercise for one hour at a time, participation in another upper limb rehabilitation intervention study, and inability to return for all study sessions. All patients gave written informed consent for participation in the study. The study was approved by the institutional review board at each center and registered with Clinicaltrials.gov (NCT02292251) prior to the start of enrollment.

### Interventions

#### Neuroanimation Therapy (NAT)

Participants played a custom-designed immersive animation-based experience: I am Dolphin (KATA, Johns Hopkins University) (Figure 1). 3D movements of the paretic arm controlled the movement of a virtual dolphin, swimming through different ocean scenes with various task goals including chasing and eating fish, eluding attacks, and performing jumps. Tasks were designed to promote movement in all planes throughout the active ranges of motion, and titrated based on successful completion of progressive levels of difficulty.

**Figure 1.**
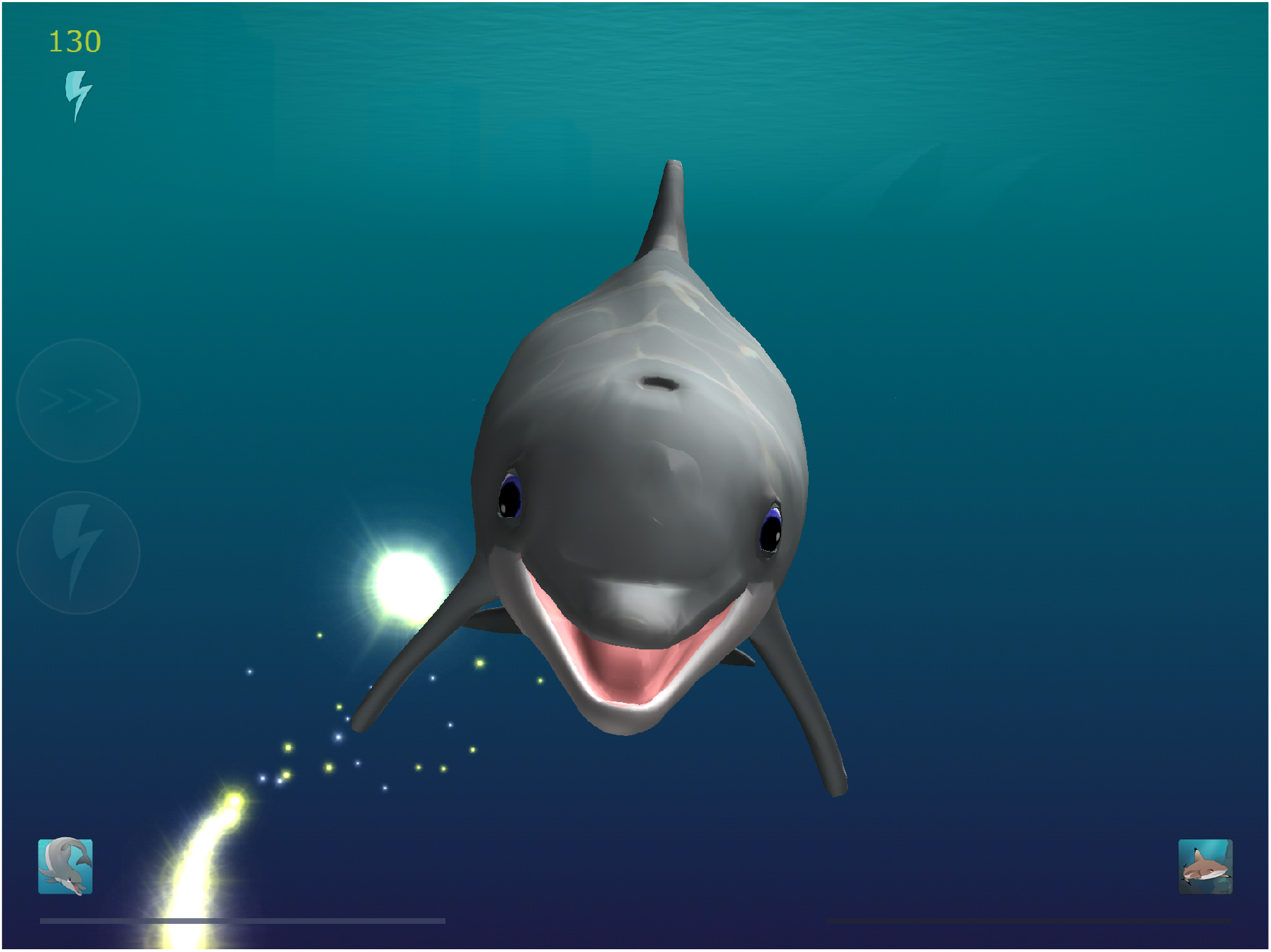
Participants in the exploratory therapy group played the MindPod Dolphin game while their arm was unweighted by the Armeo®Power exoskeleton device.

Patient’s paretic arm was unweighted using the Armeo®Power (Hocoma AG, Volketswil, Switzerland), an upper limb exoskeleton device. This allowed practice of 3D arm movements despite antigravity weakness without requiring a therapist to actively lift the paretic arm. The degree of unweighting provided by the exoskeleton was adjusted for each patient to maintain shoulder flexion to 90 degrees at rest so as to provide weight-support of the paretic limb throughout its full active range in all directions. No active assistance was given along the line of movement by the device. The device was integrated with a custom gaming software in a room which simulated an immersive oceanic environment. A large screen displayed the dolphin in his environment, oceanic sounds and music were played, and the lights were dimmed for the entirety of each session. A licensed physical or occupational therapist was present throughout each session and provided feedback to encourage normal (non-synergistic) movement patterns at all times.

#### Modified conventional occupational therapy (COT)

COT was administered by a licensed therapist according to a written standardized protocol. Active COT time was matched to that of the NAT group (60 minutes per session). The Chedoke-McMaster Stroke Assessment Scale^16^ was administered at the beginning of each week to guide interventions aligned with the specified level of function. Therapeutic exercises consisted of range of motion (stretching) and strengthening exercises of the paretic arm, and training of ADLs, such as simulated cooking/eating, dressing, grooming and cleaning tasks. A typical COT session would include 30 minutes of impairment intervention (e.g. scapular stability, weight bearing, active range of motion, stretching, and strengthening), and 30 minutes of activity training (e.g. reaching, grasping, pinching, bilateral limb coordination during functional activities, ADLs). The first session of the day targeted shoulder/elbow, the second session targeted wrist/hand. The musical soundtrack of NAT was played during COT to maintain blinding if the therapy was conducted in close proximity to blinded evaluators.

All therapeutic activities were documented by the therapy team, including the amount of assistance provided (e.g. passive mobility, active assisted mobility, weight bearing), any modifications to the exercise, and total time spent on each area of the upper limb.

### Clinical study outcomes

At the baseline visit, patients underwent a comprehensive clinical evaluation that included: medical chart and radiological review, National Institutes of Health Stroke Scale (NIHSS), proprioception assessment (using distal phalanx testing, abnormal defined as <3/5 trials incorrect), Montreal Cognitive Assessment^17^, Florida Apraxia Battery^18^, star cancellation test of visuospatial neglect (presence of hemineglect determined by cutoff score of 44^19^), and Beck Depression Inventory-II^20^. Study assessments were performed at four timepoints: baseline (pre-training), and post-training days 3 (±2 days), 90 (±10 days), and 180 (±10 days). All evaluators underwent video and in-person training prior to conducting study assessments and were blinded to patients’ treatment allocation.

The primary outcome measure was the change in upper limb impairment measured by FM-UE, from baseline to post-training day 3. The FM-UE is a widely used, reliable, and validated measure of motor impairment in patients with stroke^21^. It evaluates the ability to make upper limb movements in and out of synergy patterns and consists of 33 items graded on an ordinal scale (0-2), with a best possible score of 66. The minimal clinically important difference (MCID) for the FM-UE is considered to be approximately 10% of the maximum score, or 6.6 points^21^.

Secondary clinical outcome measures included changes from baseline to day 3 post-training in: 1) ARAT, an assessment of upper limb activity limitation with the MCID for the paretic arm being 12 points^22^ 2) grip strength using a Jamar dynamometer (average of three trials, MCID 5kg^22^), and 3) the hand domain of the Stroke Impact Scale (SIS hand) version 2.0, a self-reported measure of hand function^23^. All measures were also assessed at 90 and 180 days post-training, to evaluate longer-term gains. Measures of therapy compliance included number of sessions completed and minutes of active therapy within each session.

### Kinematics of planar reaching

Motor control of the proximal arm was evaluated using a planar arm reaching task and analyzed using methods that have been previously described^14^ (for detailed information, see Supplemental Materials).

### Finger strength and individuation

Finger strength and individuation were evaluated using an ergonomic keyboard device that measures isometric forces produced by each finger^24^ (for detailed information, see Supplemental Materials).

### Statistical analysis

Each outcome measure (FM-UE, ARAT, grip strength, SIS hand, AMD^2^, finger MVF and individuation index) was analyzed using the same framework and model structure. Specifically, we used a linear mixed model in which timepoint was treated as a categorical predictor with four levels (reference: baseline visit), therapy type was a categorical variable with two levels (reference: NAT), and all timepoint by therapy type interactions were included. Additionally, a subject-specific random intercept accounted for within-subject correlation across timepoints. This model structure estimates the mean change in outcome value from baseline to each timepoint as well as the difference in this change over time comparing NAT to COT groups, using all available subject data at each visit. Regression diagnostics were used to assess model fit, and two-sided Wald tests were used to assess statistical significance for group-level comparisons.

We conducted a power analysis for the ability to detect a difference in treatment effect between NAT and COT groups. This analysis was based on a two-sample t-test with two-sided alpha level set at 0.05 and assumed the difference between groups in the change in FM-UE score would be 7 (MCID). For an effect standard deviation of 5, 10 subjects per group yields 84% power to detect the true alternative; for an effect standard deviation of 7, 20 subjects per group yields 87% power. We planned to randomize 24 subjects in each group, based on our observed attrition rate of 20% for previous longitudinal stroke studies.

#### Comparison to historical cohort (HC)

In exploratory analyses, we sought to compare FM-UE and ARAT outcomes among patients in the current study, all of whom received an intensive intervention, to patients who received usual clinical care. To do so, we used data from the EXPLICIT trial^25^, which was conducted in the Netherlands and randomized patients within 14 days of first-ever hemiparetic stroke to modified constraint induced movement therapy or usual care. Specifically, patients in the EXPLICIT usual care cohort received occupational therapy based on current Dutch guidelines for 30 minutes a day, five days a week for three weeks beginning within five weeks post-stroke. Usual care patients from the EXPLICIT trial were matched to patients in our intense treatment groups (entire SMARTS2 cohort) based on day post-stroke (± 4 days) and severity (± 4 points for FM-UE and ARAT). After candidate matches were found, we randomly sampled a single subject to serve as the match for each subject in the intense treatment group. In this analysis, we computed the change in FM-UE and ARAT between baseline and day three post-training for patients in the SMARTS2 group and a similar timeframe for the matched patients in the EXPLICIT usual care group, and compared the average change between groups. Because there can be several potential matches of whom one is selected, we repeated the full analysis and aggregated results to account for uncertainty in the matching process.

## RESULTS

Between April 2015 and October 2017, 4030 patients with ischemic stroke were assessed for eligibility (Figure 2). The study was stopped after 24 subjects were randomized due to slow recruitment. Table 1 summarizes baseline characteristics of patients who were randomized. The median number of days from stroke onset to baseline assessment was 19.0 days (IQR 12.0, 33.0) and 14.0 (IQR 12.5, 35.5) in the NAT and COT groups, respectively. There were no significant differences in gender, arm affected, proportion receiving tissue plasminogen activator or mechanical thrombectomy, median NIHSS score, proportion with hemineglect, proportion receiving selective serotonin reuptake inhibitors, or baseline FM-UE or ARAT scores. The COT group did have statistically worse scores on the Florida Apraxia Battery, whereas the NAT group had worse depression scores at baseline.

**Figure 2.**
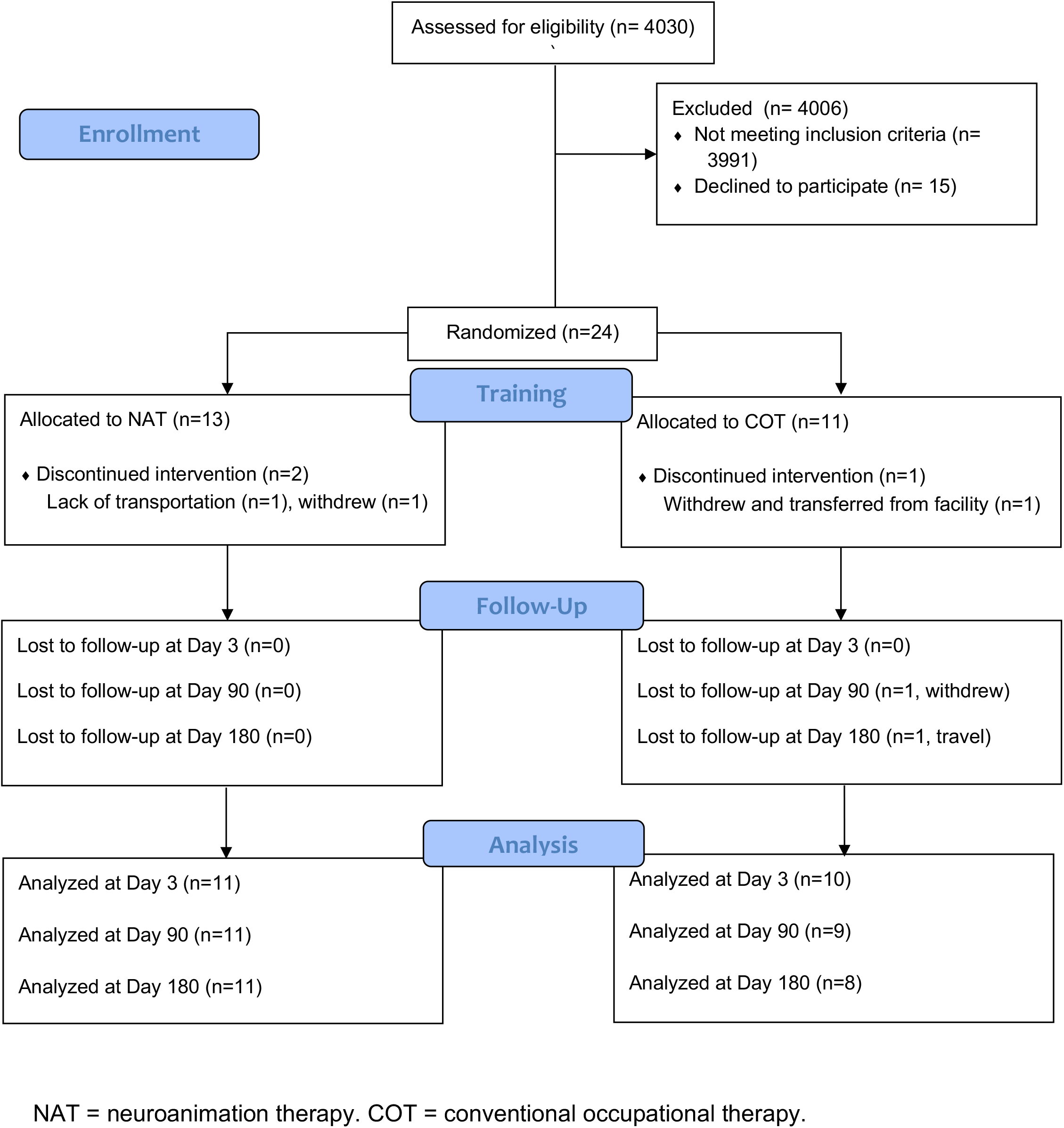
Participant flow through the study.

**Table 1.** Participant characteristics at baseline

|  | **NAT**<br>(n=13) | **COT**<br>(n=11) | **p-values** |
| --- | --- | --- | --- |
| Age in years (SD) | 62.0 (10.4) | 64.4 (14.0) | 0.640 |
| Gender male (%) | 5 (38.5%) | 6 (54.5%) | 0.706 |
| Nondominant affected | 7 (53.8%) | 5 (45.5%) | 1.000 |
| Received tPA (%) | 10 (76.9%) | 7 (63.6%) | 0.793 |
| Thrombectomy (%) | 2 (15.4%) | 3 (27.3%) | 0.834 |
| Days between stroke onset and baseline assessment (median [IQR]) | 19.0 [12.0,33.0] | 14.0 [12.5,35.5] | 0.663 |
| NIHSS (median [IQR]) | 6.0 [6.0,10.0] | 6.0 [5.5, 9.5] | 0.859 |
| Florida Apraxia Battery (median [IQR]) | 15 [15, 15] | 14 [12.5, 15] | 0.031 |
| Hemineglect (%) | 1 (7.7%) | 1 (10.0%) | 1.000 |
| Abnormal proprioception (%) | 2 (15.4%) | 5 (45.5%) | 0.244 |
| Beck Depression (median [IQR]) | 17 [11,19] | 5 [3,8] | 0.025 |
| Taking SSRI (%) | 10 (76.9%) | 4 (36.4%) | 0.111 |
| Baseline FM-UE (median [IQR]) | 23.8 (12.1) | 22.2 (8.7) | 0.562 |
| Baseline ARAT (median [IQR]) | 10.0 [3.0, 33.0] | 9.0 [3.0, 21.5] | 0.640 |

NAT = neuroanimation therapy. COT = conventional occupational therapy. tPA = tissue plasminogen activator. NIHSS = National Institutes of Health Stroke Scale. Florida Apraxia Battery scores range 0-15 with a lower score indicating worse apraxia. SSRI=selective serotonin reuptake inhibitor. FM-UE=Fugl-Meyer Upper Extremity motor score. ARAT=Action Research Arm Test

One patient in the NAT group did not receive the intervention due to transportation issues. Two patients withdrew from the study before the end of the planned intervention: one preferred to receive only standard rehabilitation, and one was transferred to another facility. Compliance with active therapy was high in both groups. Excluding the 3 patients who did not receive the intervention or withdrew before the end of therapy, the mean total time in active therapy was 1769 minutes for NAT (98% of target) and 1801 for COT (100% of target). Two patients in the NAT group required minimal assistance from the therapist to initiate movement in the first few sessions but were subsequently able to complete the training on their own. Twenty-one patients completed the post-training day 3 assessment (88%), 20 completed the post-training day 90 assessment (83%), and 19 completed the day 180 assessment (79%).

### Clinical outcomes

Our primary outcome was the change in FM-UE score from baseline (pre-training) to post-training day 3. There was no significant difference between NAT and COT groups in our primary outcome of FM-UE changes from baseline to day 3 post-training (difference 1.34, standard error [SE] 5.15, p=0.797), or in FM-UE changes from baseline to day 90 (difference -3.28, SE 5.26, p=0.316) and day 180 (difference -0.09, SE 5.38, p=0.132) post-training (Table 2). There was similarly no significant difference between NAT and COT for the change in ARAT from baseline to day 3, day 90, or day 180 post-training (Table 2). Grip strength and SIS hand also showed no between-group differences in change from baseline at any timepoint (Table 2).

**Table 2.**
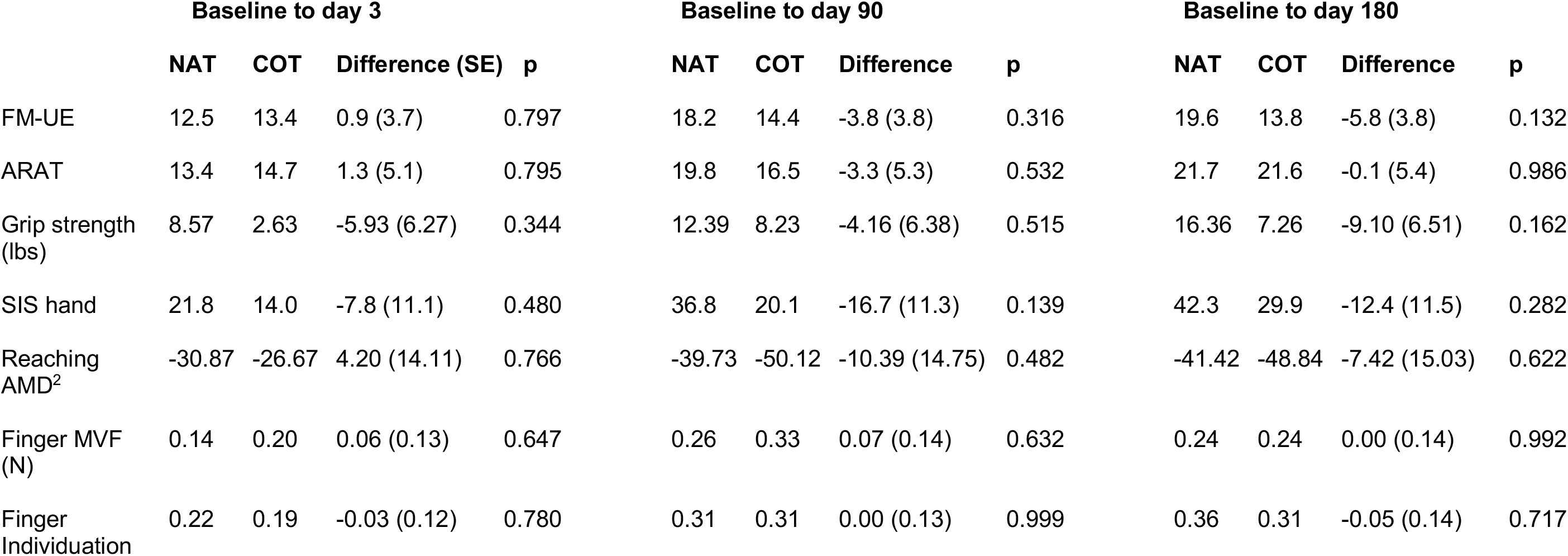
Results of mixed model estimates of changes from baseline to post-training days 3, 90, and 180. There were no significant differences between NAT and OT groups in any of the primary or secondary outcome measures. Abbreviations: NAT=neuroanimation therapy; COT=conventional occupational therapy; SE=standard error; p=p-value; FM-UE=Fugl-Meyer Upper Extremity motor assessment; ARAT=Action Research Arm Test; SIS hand=Stroke Impact Scale v.2 hand domain; AMD^2^=average squared Mahalanobis distance; MVF=maximum voluntary force.

### Reaching kinematics

We calculated the average squared Mahalanobis distance (AMD^2^) for reaching trajectories performed with the paretic arm at the four timepoints, compared to a reference population of neurologically-healthy control subjects. There were 2 patients in the COT group whose reaching kinematic data were excluded from analysis because there were too few movements in the baseline session that were suitable for inclusion in the analysis. An additional 2 patients were missing baseline assessments of reaching kinematics (1 in each group) and therefore were not included in the analysis of change scores from baseline. Of the remaining patients, day 90 data were missing for 2 subjects and day 180 data were missing for 2 patients, due to time constraints or withdrawal from the study. There was improvement in reach kinematics across the intervention period but there was no significant difference in the change in AMD^2^ from baseline between NAT and COT groups at day 3, day 90, or day 180 post-training (Table 2). There was a significant correlation between the AMD^2^ and the ARAT *(p* = 0.028) but not the FM-UE (*p* = 0.622).

### Finger strength and individuation

Maximum voluntary force and finger individuation index were calculated for the more affected arm at the four timepoints as described above. 1 patient in the NAT group did not complete the tasks at baseline and was excluded from analysis. There were also data missing for 4 patients at post-training day 90 (2 in NAT and 2 in COT), and 5 patients at post-training day 180 (2 in NAT and 3 in COT). We found no significant between-group difference in the change in MVC or individuation index from baseline to any post-training timepoint. (Table 2).

### Comparison of intensive therapy to the historical cohort (HC)

In an exploratory analysis, we compared the change in FM-UE and ARAT from baseline to post-training day 3 in our SMARTS2 study cohort (NAT and COT groups combined) to changes across a similar timeframe with usual care in a historical cohort from the EXPLICIT-stroke study^25^. Patients in SMARTS2 were matched by time post-stroke and severity (FM-UE and ARAT) with patients from EXPLICIT. We observed a significant benefit in upper limb activity (ARAT difference 7.33, SE 2.88 pts, p=0.011) but not for impairment (FM-UE difference 1.44, SE 2.57, p=0.564) with the intensive therapy provided in SMARTS2 compared with usual care (Figure 3).

**Figure 3.**
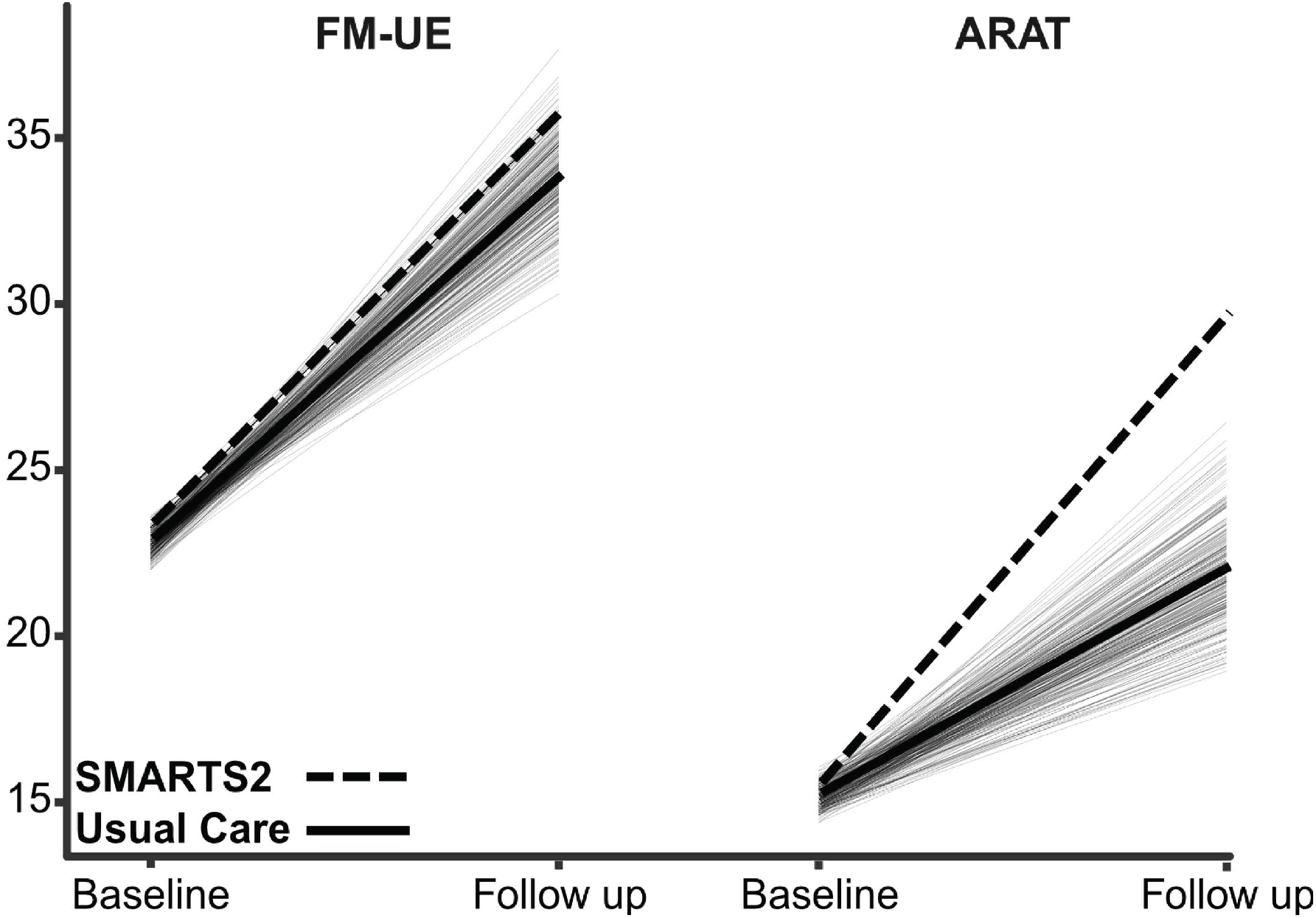
Comparison between the SMARTS2 study cohort receiving intensive therapy (dotted line) and a historical cohort from the EXPLICIT trial receiving usual care (means represented by heavy lines, individual subjects by thin solid lines). The groups were matched for baseline time post-stroke and severity. A significant benefit in upper limb activity and dexterity (ARAT), but not for upper limb impairment (FM-UE), was seen with the intensive therapy provided in SMARTS2.

### Adverse events

There were a total of 55 adverse events (AEs) that occurred in 13 patients during the study. There were 4 serious adverse events in the COT group that were unrelated to the study procedures, 2 in the NAT group (2 fall resulting hospitalization and 2 unrelated medical condition) Of the 51 non-serious AEs, 23 (42%) occurred in the NAT group and 32 (58%) occurred in the NAT group and 7 (23%) occurred in the COT group. In the NAT group, 5 AEs were probably related (neck pain, fatigue in 3 patients, and bruising) and 6 AEs were possibly related (eye pain in 2 patients, headache, nausea, worsened ataxia, and a fall) to study procedures. In the COT group, two AEs were definitely related (wrist pain in 2 sessions), 1 probably related (pain), and two possibly related (pain, fall) to study procedures.

## DISCUSSION

In this randomized, single-blinded proof-of-concept trial we sought to test the idea, inspired by studies in non-human primates, that high-intensity and high-dose upper limb therapy focused on movement quality rather than task accomplishment, and delivered early after stroke, would reduce motor impairment more than usual care does. We tested this main idea by taking two distinct approaches. The first was administering high doses of conventional upper limb therapy. The second was a new immersive animated experience that centers on a proprietary form of animation designed to promote playful exploration of high quality continuous 3D arm movements^15^. Here we found that both approaches led to similar changes in the FM-UE, ARAT, reaching kinematics, finger strength, and finger individuation. Looking at a historical cohort, we found that both approaches were superior to usual care with respect to the ARAT but not the FM-UE. Unfortunately, hand and planar kinematic measures were not available for the historical cohort.

Conventional occupational therapy (COT) mainly emphasizes repetitive task-oriented training (TOT)^26,27^, an approach predicated on practice schedules based on motor learning principles. Not much time is dedicated to the upper limb in regular therapy sessions^12^, which means that TOT focuses more on compensatory movements for task accomplishment. That said, there is nothing inherent to COT that precludes a switch in emphasis to movement quality, especially if therapists are given more time with the patient, as they were in SMARTS2. In three recent studies in patients with chronic stroke, large gains in both the ARAT (or other activity-level measure) and the FM-UE were seen when patients were provided with either five or six hours of upper limb therapy a day for five days/week for three, six or twelve weeks^3-5^. Clearly these are very high intensities and doses of therapy. In two of the studies^4,5^, the authors explicitly state that they wanted to make “movement practice as close to normal as possible”, and did so by progressing from single-joint to two-joint movements, then assembling these into task components and finally practicing performance of the full task. It is evident that they combined the more traditional neurophysiological approach, which focuses on movement quality, with TOT. The therapists in our study took a comparable approach in the COT group, as outlined in the methods section. In fact, they explicitly stated on questioning that they were able to focus on movement quality precisely because they had more time with the patient.

The NAT and COT groups showed comparable changes in ARAT scores. These changes were significantly greater than the changes seen in the historical control group receiving usual care over the same time period. The ARAT is a valid and responsive measure of upper limb activity on the ICF scale^28,29^ and its components have considerable overlap with the reach and grasp tasks in non-human primate experiments investigating motor recovery. Indeed, the ARAT correlates well with kinematic measures of reach and grasp^30^.

We argue here that the changes in the ARAT in the NAT and COT groups are an indication of true improvement in the quality of arm and hand motor control and not just compensation, even though performance on items scored less than 3 (i.e., “normal”) can include compensatory movements. First, we know that the ARAT can show, just like the FM-UE, changes, as we saw here (13.4 and 14.70 points from baseline to day 3 post-training in NAT and COT groups, respectively), that are larger earlier compared to later after stroke ^31^. That ARAT changes are greater when high-dose therapy is given earlier than later means suggests that they, at least in part, reflect true restitution and not just learned compensation. Second, if the two intervention groups were just being trained to compensate better than usual care, then this must be because they learned to compensate *during* the intervention. This would not be possible for the NAT group, however, because there was no prehension training of any kind – the virtual dolphins were steered with the arm only. Third, large improvements in activity measures have been seen with intense and high dose COT in patients with chronic stroke^5,6^. These changes seem to be dose-dependent, only becoming large when 90 hours or more of treatment are given^3-5^. In contrast, 32 hours does not lead to large ARAT changes^31^. If large ARAT changes were just due to optimizing compensatory strategies it seems unlikely that they would not also be seen after 32 hours of COT. Finally, here we found a significant correlation, as have others^30,32^, between improvement in the quality of arm kinematics and the ARAT. Thus, like in non-human primates^8^, we conclude that intense and high-dose upper limb therapy focused on movement quality can improve the control of prehension movements in the subacute period after stroke.

Notably, we did not see an increase in the FM-UE, our primary outcome measure, beyond what was seen in the usual care historical control group. Of course, as we were studying subacute rather than chronic stroke, there were large changes in the FM-UE due to spontaneous recovery but we were not able to augment them with either of our interventions. It is always possible that this is a false negative result given the low *n*, but a previous study of early intense and high-dose upper limb therapy, in this case constraint-induced movement therapy, also reported a dissociation between the ARAT and the FM-UE^25^.

It is possible for ARAT improvements to reflect true changes in motor control and yet not be detected by the FM-UE score due to differences in emphasis for the two scales. For example, a patient who has regained active range of movement in the shoulder/elbow but has persistent difficulty with out of synergy movements (e.g., unable to initiate shoulder flexion or abduction without bending the elbow, which would confer a score of “0” on these FM-UE items) may nevertheless improve on the ARAT by gaining the ability to reach the top shelf through improved shoulder flexion and elbow extension. Another situation in which one may see a dissociation between FM-UE and ARAT changes is a patient who primarily regains distal dexterity, which is weighted more heavily on the ARAT than the FM-UE.

The differential responsiveness of the FM-UE and the ARAT to an intervention is not altogether surprising. FM-UE and ARAT measure different constructs of the ICF model, reflecting the levels of body function and activity, respectively. In addition, the FM-UE scale was primarily devised to quantify post-stroke synergies over the course of motor recovery ^33^ whereas the ARAT emphasizes assessment of prehension during more functional tasks^29,34^. In other words, one primarily targets a positive sign and the other a negative sign of the upper motor neuron syndrome^8^. That being said, synergies will intrude on a functional task, especially in the absence of arm weight support^35,36^, which is why the two measures often correlate with each other^37,38^. It is unfortunate that the FM-UE has come to be considered synonymous with overall impairment after stroke, even though it was designed to assess mostly a single component of impairment, namely synergies, over strength or dexterity. This is problematic because thus far it seems that the positive and negative symptoms of stroke respond differently to interventions in the sub-acute period. In this study, we saw dissociation between the ARAT and the FM-UE, and a similar finding was reported when extra sessions of constraint-induced movement therapy were added in the sub-acute period^25^. It is to be hoped that new kinematic measures will soon be developed that can distinguish arm dexterity/quality of motor control from both synergies and compensation during performance of 3D functional tasks^39,40^. This is of utmost importance because hemiparesis in humans appears to be both a deficit disorder related to damage to the corticospinal tract^41,42^ and a movement disorder, perhaps related to upregulation of the reticulospinal tract^43-45^. These positive and negative signs of hemiparesis will likely need distinct forms of intervention.

Our findings for the NAT and COT groups are congruent with what has been reported in many recent neurorehabilitation studies and trials – both the novel intervention group and the control group show similar, and often large, treatment responses. This has been taken as evidence that new technological or pharmacological interventions do not outperform higher intensities and doses of conventional therapy or, by extension, usual clinical care. In a recent review of 15 neurorehabilitation trials conducted in the last five years it is stated: “There is no clear evidence that interventions tested in large multicenter stroke rehabilitation trials are superior to current care. Furthermore, patients benefited from both the experimental and control interventions at both the subacute and chronic stages”^46^. The crucial point being missed here, however, is that control interventions in clinical trials often consist of *more*, and sometimes considerably more, conventional therapy than is usually given during regular clinical care. This is certainly the case in our study where patients in the COT group received two hours a day of therapy for five consecutive days over three weeks. In addition, trials tend to select for patients with less comorbidity and fewer cognitive deficits, which allows them to receive higher doses of usual care. Therefore, the positive results for controls in trials do *not* imply that usual conventional care is equally efficacious to the novel intervention. Indeed, trials that have directly compared higher doses with usual doses of conventional care have found a difference between them^47-49^. Thus, the fact that a new approach, like the NAT here, is as good as high doses of COT should be taken as a reason for optimism. The equivalence implies that the new intervention must possess an active ingredient that potentially could be further optimized in terms of efficacy, efficiency, cost-effectiveness, and scalability. This situation can be considered analogous to what happened in the evolution of thrombectomy for acute stroke. In 2013, three trials showed no benefit of thrombectomy over usual care^50-52^. By 2015, five trials showed superiority for thrombectomy^53^. What happened? The two main reasons were choice of the correct technology for clot removal and a change in protocol design.

As with post-stroke thrombolysis, it is possible that we are on the cusp of a change in the delivery and efficacy of upper limb neurorehabilitation. In this case, the correct choice would be to move toward more immersive experiences to promote intense exploratory training with a focus on movement quality. The protocol change would be, as in SMARTS2, to encourage exploratory multi-joint movements outside of a task context for at least two hours a day. Thus, based on the results here, we suggest that an alternative to just increasing the amount of time available to administer COT, is to devise technology-based solutions that make it easier and more enjoyable to deliver higher doses and intensities of impairment-focused therapy^54^. It should also be emphasized, that even though in this small study we found equivalence between COT and NAT for delivery of higher doses of intense upper limb therapy, it does not need to be either/or. It may turn out that the two approaches can complement each other. COT could be considered analogous to drills in sport, for example practicing backhands in tennis for an hour. NAT could be the holistic approach where you combine all the components into a full game.

Compliance with therapy was high in both NAT and COT groups, reaching 98% and 100% of targeted time on task, respectively. Overall both interventions were safe, with no serious adverse events related to study procedures. There were, however, more adverse events in the COT group than the NAT group. Fatigue was reported more often in the NAT group, which therapists did not always perceive to be a negative because the therapy was designed to be challenging. Other side effects in the NAT group such as transient headache and pain have been reported previously with game-based interventions and are not unexpected with high intensity training^55^.

This study clearly has a number of limitations. First, the number of patients in this proof-of-concept trial was low. Indeed, we recruited only half the number of patients we anticipated. This is attributable both to our inclusion/exclusion criteria and the challenges of providing two hours of time-on-task upper limb therapy, five days a week beginning in the first six weeks after stroke in addition to usual care^56,57^. Second, we were only able to begin our interventions after patients were discharged from in-patient acute rehabilitation at the sites in the United States due to the challenges of delivering high doses of therapy in addition to standard care in the inpatient rehabilitation setting as well as the need for short length of stay^58^; the average start time was therefore about three weeks post-stroke. From our previous work, we have shown that the time window of heightened neuroplasticity responsible for spontaneous recovery, and perhaps for enhanced training-related improvement that takes advantage of this heightened neuroplasticity, might be as short as 5 weeks^14^. This difficulty with our enrollment time window attests to the continuing challenge of conducting neurorehabilitation trials in this early time period after stroke^46,58,59^. Third, we had to use a historical usual care group, albeit an extensive and well-matched one^25^. This was necessary because we were not able to ask patients to enroll in a trial offering three weeks of extra care with the chance that they would end up in the control group that got no extra therapy but would nevertheless require them to make trips to the hospital for assessment. Given our strict time window offering the active intervention later was not an option. Another point, as we made above, is that in clinical trials, the control intervention is most often not “usual” care but an amplified and often unrealistic version of it^46^. Here we were fortunate that a cohort existed that delivered care of the upper limb that was close to what patients actually receive in the subacute post-stroke period.

## CONCLUSIONS

Increasing the dose and intensity of upper limb rehabilitation training early after stroke, with focus on movement quality, led to gains beyond those seen with usual care. This additional improvement was achieved either by having therapists provide much more COT or with a novel exploratory animation-based approach with exoskeletal weight support. This equivalence is exciting, as it suggests that an immersive animation-based experience combined with weight-support might pave the way forward for providing high doses of upper limb rehabilitation focused on movement quality in a more efficient, enjoyable, and scalable way at any time post-stroke.

## Data Availability

The datasets generated during and/or analyzed during the current study are available from the corresponding author on reasonable request.

## ACKNOWLEDGMENTS

This study was principally funded by the James S. McDonnell Foundation Grant 220020220 (J.W.K.). Additional funding came from the P&K Pühringer Foundation (A.R.L.), Sheikh Khalifa Stroke Institute (SKSI) (J.W.K., O.A., P.R., R.S.) and the Neurology and PMR Departments at the Johns Hopkins University School of Medicine. J.G.’s work was supported in part by NIH grant R01NS097423. The EXPLICIT-stroke consortium and the EXPLICIT-stroke cohort were funded by The Netherlands Organization for Health Research and Development (ZonMw Grant No. 89000001). JWK, OA, and PR are co-inventors of the underlying dolphin technology, which has now been licensed to MSquare Healthcare (a MindMaze Inc. Company) and is available as an approved device called MindPod Dolphin.

